# Neuroscience-Informed Classification of Prevention Interventions in Substance Use Disorders: An RDoC-based Approach

**DOI:** 10.1101/2022.09.28.22280342

**Authors:** Tara Rezapour, Parnian Rafei, Alex Baldacchino, Patricia J. Conrod, Geert Dom, Diana H. Fishbein, Atefeh Kazemi, Vincent Hendriks, Nicola Newton, Nathaniel R. Riggs, Lindsay M. Squeglia, Maree Teesson, Jasmin Vassileva, Antonio Verdejo-Garcia, Hamed Ekhtiari

## Abstract

Neuroscience has contributed to uncover the mechanisms underpinning substance use disorders (SUD). The next frontier is to leverage these mechanisms as active targets to create more effective interventions for SUD treatment and prevention. Recent large-scale cohort studies from early childhood are generating multiple levels of neuroscience-based information with the potential to inform the development and refinement of future preventive strategies. However, there are still no available well-recognized frameworks to guide the integration of these multi-level datasets into prevention interventions. The Research Domain Criteria (RDoC) provides a neuroscience-based multi-system framework that is well suited to facilitate translation of neurobiological mechanisms into behavioral domains amenable to preventative interventions. We propose a novel RDoC-based framework for prevention science and adapted the framework for the existing preventive interventions. From a systematic review of randomized controlled trials using a person-centered drug/alcohol preventive approach for adolescents, we identified 22 unique preventive interventions. By teasing apart these 22 interventions into the RDoC domains, we proposed distinct neurocognitive trajectories which have been recognized as precursors or risk factors for SUDs, to be targeted, engaged and modified for effective addiction prevention.

## INTRODUCTION

Substance use disorder (SUD) is multifactorial in etiology and numerous risk factors have been implicated in its formation and progression, particularly during adolescence. At the level of prevention, several approaches have been proposed to target some of these factors through educational and socio-emotional skills training programs, starting from early childhood (e.g., Promoting Alternative Thinking Strategies; PATHS) (Riggs et al., 2006). Programs that are largely focused on adolescents in school settings tend to harness social and behavioral theoretical models such as the social influence model, the social learning theory, and the theory of planned behavior (Kempf et al, 2017).

These programs are mainly embedded within the educational structure and include content to increase adolescents’ awareness of substance use related harms and various social influences, to correct inaccurate adolescents’ perception regarding the prevalence and popularity of SUD, and to teach life skills (e.g., problem-solving, decision-making skills) (Griffin & Botvin, 2010). Building from these models, programs such as PREVENTURE (Conrad, 2016), CLIMATE schools (now called OurFutures) (Slade et al., 2021), Life Skills Training (LST) (Botvin et al., 1990), and keepin’ it REAL (Kulis et al., 2007) have been developed, implemented and found to have an acceptable degree of efficacy (Tremblay et al., 2020).

Over the past few decades, however, our understanding of SUD has been reshaped by the evidence from neuroscience suggesting SUD can be characterized by certain functional indicators that transcend traditional diagnostic boundaries and act as pre-diagnostic markers that could be targeted through preventive approaches (Debenham et al., 2021; Fishbein et al., 2016). Developmental neuroscience informs us that during adolescence, the development of different brain structures occurs at various rates. The structures (i.e., limbic regions) that are implicated in emotional processes undergo early maturation, while those involved in executive control (i.e., prefrontal cortex) have protracted maturation (Rezapour et al., 2021). This neuroscience-informed understanding introduces adolescence as a distinct developmental stage which offers multiple opportunities to intervene on the early precursors of substance use behaviors. For example, a new personality-targeted prevention approach has emerged from the neuroscience literature which involves prophylactically intervening around psychological risk factors for early onset psychopathologies and substance use and has been shown to have beneficial effects on a broader set of outcomes compared to traditional social learning-based prevention programs (Newton et al., 2021).

Additionally, numerous studies have found that variation in several neuropsychological functions plays a role in different stages of SUD. Current neuroscience-based models (Koob & Volkow, 2016; Yücel et al., 2019) conceptualize SUD as neuroadaptive / neurodevelopmental processes that happen at two different time scales: (1) a recurring cycle of binge/intoxication, withdrawal/negative affect, and preoccupation/anticipation (craving) stages; and (2), a protracted “allostasis” that progressively alters neurotransmitter and stress responses, resulting in neuroplastic changes in brain reward, stress, and executive function systems. Identifying the neurocognitive domains implicated in each stage has considerable potential to help practitioners and clinicians improve their insight into SUD and apply that knowledge to more effectively treat and/or prevent SUD (Ekhtiari et al., 2021; Meredith et al., 2021; Debenham et al., 2020). Additional conceptualizations of SUD have focused on neurodevelopmental processes (Rose et al., 2019; Conrod and Nikolaou, 2016) to highlight the importance of individual differences and contextual factors such as trauma (Laroque et al., 2022), in moderating the above processes in the formation of SUD (Morin et al., 2018; Afzali et al., 2017; 2021). However, a comprehensive neuroscience-based conceptual framework that could inform underlying neurobiological mechanisms in SUD development is still lacking to guide effective design of preventive interventions.

In 2010, the National Institute on Mental Health (NIMH) launched the Research Domain Criteria (RDoC) as part of its strategic plan to provide a research framework for studying psychiatric disorders, including SUDs (Insel et al., 2010). Grounded in neuroscience, the RDoC covers five domains: Negative Valence Systems, Positive Valence Systems, Cognitive Systems, Systems for Social Processes, and Arousal and Regulatory Systems. There is a new suggestion to add a new domain to RDoC to cover sensorimotor processes (domain 6). The RDoC framework is mapped into various clinical contexts and multiple variants have been adapted including in SUD. For example, the National Institute on Alcohol Abuse and Alcoholism (NIAAA) proposed the *Alcohol and Addiction Research Domain Criteria (AARDoC)*, indexing three research domains relevant to SUD: Negative Emotionality (mapping on NIMH’s negative valence system), Incentive Salience (mapping on NIMH’s positive valence system), and Executive Function (mapping on NIMH’s cognitive system) (Witkiewitz et al., 2019) (Figure 1b). Subsequently, the *Addictions Neuroclinical Assessment (ANA*) framework was proposed to probe these domains by combining clinical, personality, genetic, neurocognitive, and neuroimaging approaches (Kwako et al., 2016). The three ANA domains are: (1) *Executive Function (including planning, working memory, attention, response inhibition, decision-making, set-shifting, and cognitive flexibility)*, associated with reduced prefrontal cortex (PFC)-mediated top-down impulse control, characterizing the preoccupation/anticipation (‘craving’) stage of the addiction cycle; (2) *Incentive Salience*, associated with phasic dopaminergic activation in the basal ganglia and the binge-intoxication stage; and (3) *Negative Emotionality (including dysphoria, anhedonia, alexithymia, and anxiety)*, associated with the engagement of brain stress systems and the withdrawal/negative affect stage of addiction. NIDA recently expanded these ANA domains and proposes NIDA Phenotyping Assessment Battery (PhAB) framework by adding two additional domains relevant to SUD (Keyser-Marcus et al., 2021; Ramey & Regier, 2019): *social cognition* (metacognition, theory of mind) and *precognition* (interoception, implicit processes, sleep), which map on NIMH’s RDoC domains of Social Processes and Arousal and Regulatory Systems, respectively (Figure 1c). The NIDA PhAB framework was developed as a “fingerprint” for addiction phenotype covering six neurofunctional domains including Metacognition, Interoception, Cognition/Executive Function, Reward/Incentive Salience, Emotion/Negative Emotionality, and Sleep/Circadian Rhythm. Due to the transdiagnostic nature of cognitive impairments in SUD, the expansion of the PhAB was suggested to include both precede (precognition) and supersede (social cognition) for potential therapeutic interventions (Figure 1d). The original RDoC framework has also been studied extensively in SUD. A Delphi study conducted by a group of addiction experts revealed a high degree of consensus on the most important components for SUD, identifying two RDoC domains (Positive Valence System and Cognitive System) and one expert-initiated construct (Compulsivity) as primary components (Yücel et al., 2019).

**Figure 1.**
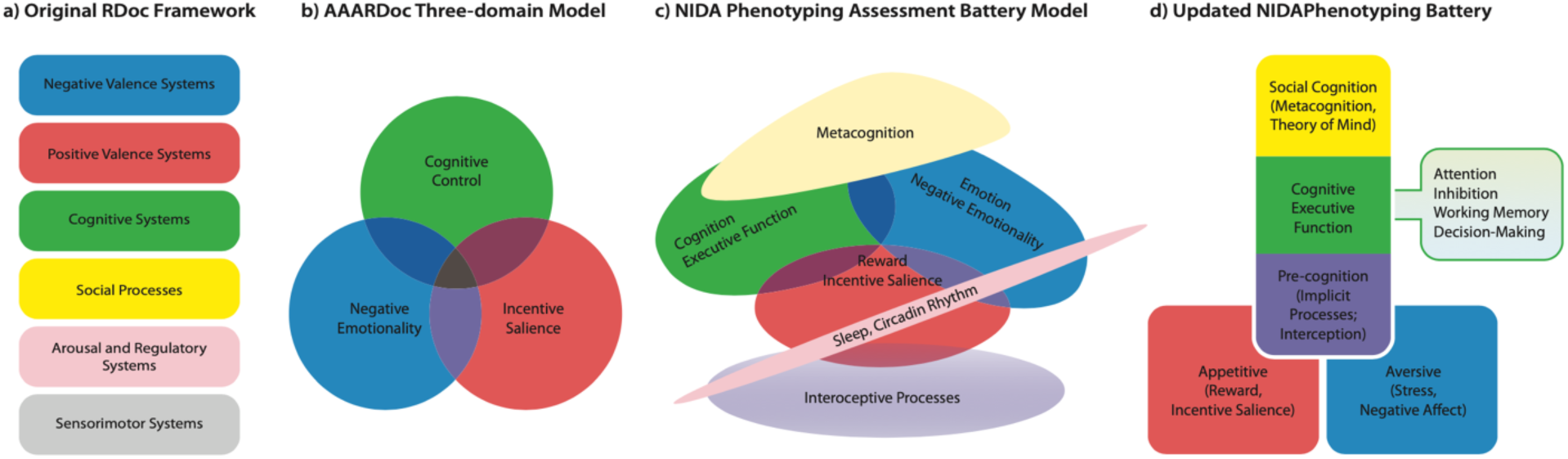
– RDoC based Addiction-related neurofunctional domains. a) The original RDoC framework includes five domains of Negative Valence System, Positive Valence System, Cognitive System, Social Processes, and Arousal and Regulatory Systems. b) The Alcohol and Addiction RDoC (AARDoC) model and the Addictions Neuroclinical Assessments (ANA) battery to assess the three-domain model, where neurofunctional abnormalities in SUDs are indexed by the three domains of Negative Emotionality, Incentive Salience, and Executive Function. c) The NIDA Phenotyping Assessment Battery (PhAB) that is designed to be administered as a set of tools to characterize “core” addiction-relevant domains in a harmonized way, for instance, across NIDA clinical trials. Interoception, Metacognition, and Sleep/circadian rhythm domains have been added to the three-domain model using a Delphi method. d) The updated NIDA Phenotyping battery comprises three transdiagnostic research domains with relevance for addiction: Appetitive motivational states (including the RDoC domain of incentive salience), Aversive motivational states (including the RDoC domain of negative emotionality), and the RDoC domain of Cognitive Executive function while includes precognition and social cognition domains.

Thus far, the interest in using neuroscience-informed models has been mainly in the context of diagnosis and targeted treatment of SUD, while there is no published framework based on the RDoC for SUD prevention (Verdejo-Garcia et al., 2019). To address this gap, the goal of this paper is to introduce an RDoC-based framework for SUD prevention. We propose a neuroscience-based model that provides a framework to identify potential precursors or risk factors for SUD and delineate mechanisms that underlie the effects of preventive interventions designed to target these factors. Based on this framework, we conducted a systematic review of school-based SUD prevention trials to identify available evidence-based interventions. The neuroscience-informed RDoC approach is then used to classify these SUD preventive interventions and their components based on their targeted RDoC domains. Such classification would increase our understanding of the key elements and neural mediators of different prevention programs and may enable their further refinement and optimization by identifying their most potent components. This approach, in turn, may indicate a potential for interfacing them with other intervention modalities targeting the same domains and personalizing them to individual or subtype needs. Therefore, by using the RDoC framework, preventive interventions could be developed not only to benefit the general population (universal prevention), but also to affect adolescents who are at risk in each domain of RDoC (selective prevention).

## RISK FACTORS FOR SUBSTANCE USE DISORDERS THROUGH THE LENSE OF RDoC

In this section, we describe the main RDoC domains that are potentially involved in SUD development and discuss how their dysfunction could increase SUD vulnerability, especially in adolescents. Figure 2 displays a model that illustrates how these domains could be considered as precursors to or risk factors for SUD development due to their non-adaptive functions in response to various stressors (a), and vice versa how they could be adjusted to protect adolescents against these stressors (b).

**Figure 2:**
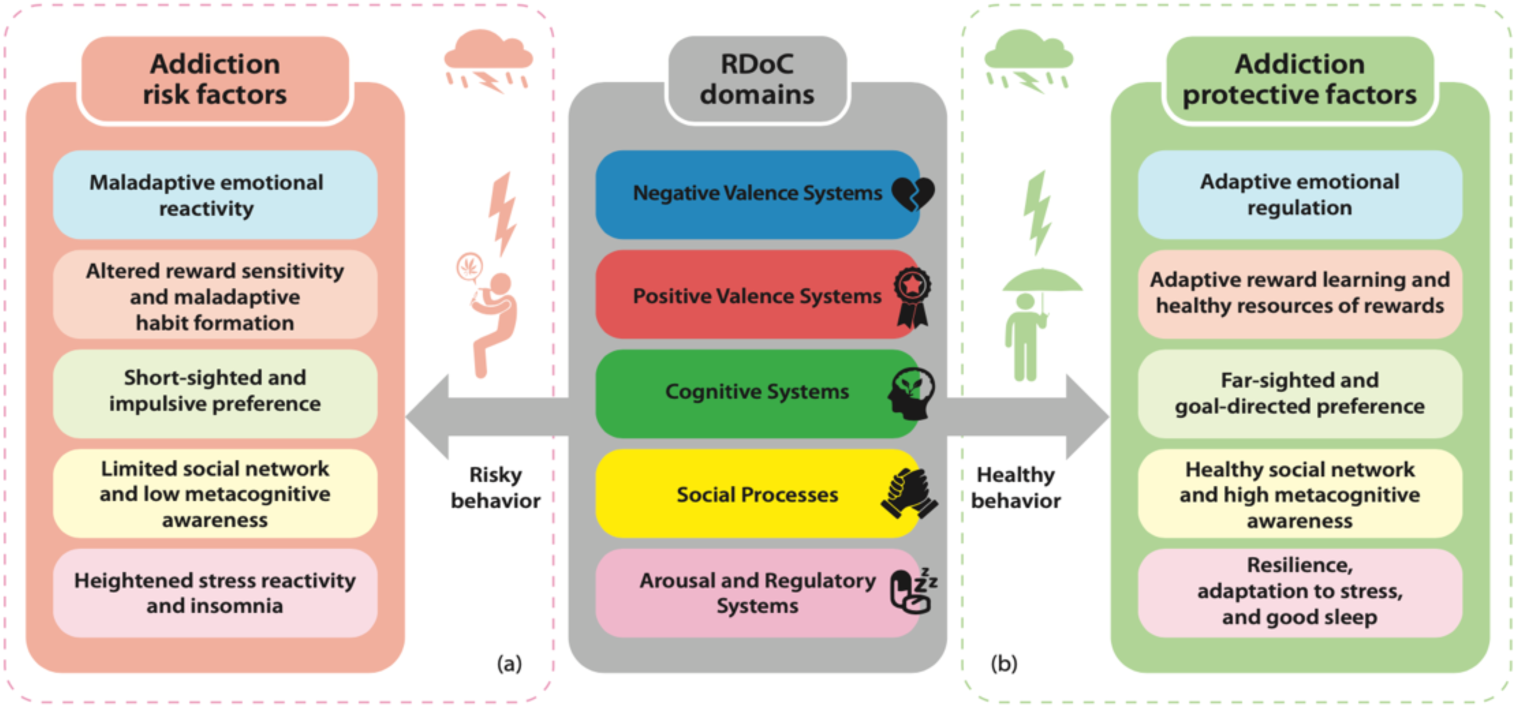
The five major RDoC domains could act as, (a) risk factors, or (b) protective factors for substance use disorders during adolescence.

Although each domain seems to be independent of the others, the previous studies reveal functional interactions between them through highly integrated neural mechanisms (Ford et al., 2014). For example, affective valence (including negative and positive) could interact with cognitive control from the domain of the cognitive system or interoceptive signals from the domain of arousal and regulatory systems (Hadley et al., 2019).

### Negative Valence Systems (NVS)

NVS is expressed in negative emotional responses (including fear, anxiety, avoidance, frustrative non-reward, deprivation of motivationally significant possession) to a particular environmental event (acute threat, ambiguous harm, prolonged threat, withdrawal of reward, loss) (Watson et al., 2017) and the brain regions that have most consistently been associated with these mental processes are the amygdala and anterior insular cortex (Büchel, 2000; Wu et al., 2014). The link between NVS and the development of SUD could be explained by the ability to regulate negative emotion in terms of both intensity and valence (Guinle & Sinha, 2020; Ohannessian & Hesselbrock, 2008).

Subjective distress can be observed as negative emotions in response to potentially aversive stimuli which then place an individual at risk for substance-seeking behaviors and craving (Zambrano-Vazquez et al., 2017). In fact, individuals who engage in substance misuse commonly exhibit maladaptive coping strategies for distress (e.g., anxiety) and often seek out the rewarding properties of abusable substances to reduce negative affect (Brooks et al., 2017).

Increased risk of SUD during adolescence is likely due in part to vulnerability to various emotionally laden challenges (e.g., romantic break-up, academic pressure, peer rejection) that increase emotional reactivity (Houck, Barker, et al., 2016; Thatcher & Clark, 2008). Limited capacity to regulate negative emotions during adolescence as a function of less connectivity between the PFC and affective limbic structures than in adulthood may result in maladaptive external regulatory strategies that place adolescents at heightened risk for SUD (Tottenham & Galvan, 2016).

### Positive Valence Systems (PVS)

The PVS include processes involved in the valuation, responding, maintaining, and learning of rewarding experiences (Swope et al., 2020). This domain is divided into several constructs, including approach motivation (motivation to obtain reward), initial reward responsiveness (hedonic responses during consummation of rewards), sustained reward responsiveness (duration of hedonic response following obtaining rewards), reward learning (linking between information about stimulus and hedonic response), and habit formation (Olino, 2016). These constructs engage a common set of brain regions in the dopaminergic system that are related to SUDs, including the ventral striatum (nucleus accumbens), orbitofrontal cortex (OFC), and anterior cingulate cortex (ACC) (Richards et al., 2013). Additional regions such as the thalamus, amygdala, insula, and inferior frontal gyrus (IFG) have also been implicated in reward processing, which often contributes to substance-seeking behaviors attributed to altered reward sensitivities (Balodis & Potenza, 2015; Silverman et al., 2015).

A potential link between PVS and SUD development in adolescents has been suggested in terms of altered sensitivity to rewarding, novel, and exciting stimuli that affects decision making (Balogh et al., 2013; Walker et al., 2017). Across development, and specifically during adolescence, increased reward-seeking behaviors, either as a result of hypo-(based on the reward deficiency hypothesis) (Cservenka et al., 2013) or hyper-responsivity of the reward system, increase the likelihood of SUD (Galván, 2010; Hardin & Ernst, 2009). Based on such explanations, adolescents place a higher value on substance use and so expect greater pleasure derived from substance use (Peeters et al., 2017). Inability to regulate responsiveness to rewards and positive emotions is a potential link between PVS and SUD initiation (Castellanos-Ryan et al., 2014; 2016).

### Cognitive System (CS)

The domain of CS encompasses a broad range of cognitive processes, including perception, attention, working memory, declarative memory, cognitive control, and language, to select, recognize, and process information to be used in goal-directed actions and future decision-making (Glenn et al., 2018). Adolescence is characterized by the asynchronous development of frontostriatal circuitry, with an impulsive striatal and affective amygdala system maturing early and being disproportionately active relative to later-maturing top–down cognitive control systems mediated by the prefrontal cortex (PFC) (Blakemore & Robbins, 2012; Casey et al., 2005; Galvan, 2010). The temporal variation of CS maturation enhances the influence of reward and emotional systems and contributes to impulsive and disinhibited behaviors, including substance use (Rose et al., 2019; Wetherill & Tapert, 2013). Several studies on adolescents indicate a link between poor executive function (i.e., inhibition, working memory) and early initiation of alcohol and other substance use (Gray & Squeglia, 2018), in line with theories such as the Reinforcer Pathology Theory (RPT) (Bickel & Athamneh, 2020). The RPT states that the value of immediate, intense, and certain addictive reinforcers (i.e., substance) would increase, whereas the value of the delayed negative outcomes and prosocial reinforcers (which are less intense and reliable) would decrease as a result of one’s short temporal window (the temporal distance over which future outcomes are considered and incorporated into present decisions and behaviors). Although such cognitive weaknesses are mainly attributed to the delayed maturation of cognitive control brain structures in adolescence, some studies support the role of family history of SUD in alcohol and early onset substance use initiation in offspring due to weaker neural connectivity (Squeglia & Cservenka, 2017; Morin et al., 2018). Overall, poor performance of the CS reduces the regulatory capacity to control socioemotional functioning and increases SUD vulnerability.

### Arousal and Regulatory Systems (ARS)

The ARS construct reflects responsiveness to internal and external stimuli, and is associated with arousal, circadian rhythms, and sleep-wakefulness (Koudys et al., 2019). The ARS also plays an important role in maintaining bodily homeostasis by using body-related information (interoceptive signals) to predict future body states and select proper approach or avoidance action (Victor et al., 2018). The hypothalamic-thalamic circuitry mainly corresponds to the regulatory systems. Also, neurocircuits related to sleep and arousal have reciprocal connections from the amygdala to other limbic structures such as the thalamus and hypothalamus, as well as to cortical structures (Henje Blom et al., 2014).

In the early course of adolescence, dysregulated stress responses (resulting from biased cognitive processes, a history of trauma, or genetic factors), combined with altered hypothalamic-pituitary-adrenal axis (HPA) axis and sympathetic nervous system responses, increases the risk of SUD development (al’Absi, 2018; Chaplin et al., 2018), particularly the misuse of substances with arousal and fear-reducing properties (Stewart, et al., 2021). In addition to the role of sleep deprivation as a stressor that triggers stress reactivity, there are several studies supporting the relationship between sleep and circadian changes and substance use in adolescents (Logan et al., 2018). Sleep problems, including circadian misalignment, sleep disturbance, and sleep loss, could affect reward systems in a way that young people are more prone toward sensation-seeking and impulsive behaviors, and thus increase the risk of substance use and risky behaviors (Spear, 2011). The negative effect of sleep problems on self-regulatory functions has been previously reported in both laboratory and field studies in adolescents (Baum et al., 2014; Louca & Short, 2014).

### Social Processes (SP)

Broadly defined, SP comprises processes and knowledge that mediate the perception and understanding of the self and others, as well as the responses that are generated within a social context (reception and production of facial and non-facial communication) (Koudys et al., 2019). A recent meta-analysis used the activation likelihood estimate method and reported that the medial prefrontal cortex (mPFC), anterior cingulate cortex (ACC), posterior cingulate cortex (PCC), temporoparietal junction (TPJ), bilateral insula, amygdala, fusiform gyrus, precuneus, and thalamus are the neural underpinnings of the SP domain (Lobo et al., 2022).

To explain how this system contributes to SUD development during adolescence, we refer to the role of metacognition (self-knowledge) in the context of within-person characteristics (e.g., inaccurate emotional awareness, self-efficacy) and the role of affiliation and attachment in the context of between-person interactions (e.g., normative misperceptions) (dos Santos Kawata et al., 2021; Shadur & Hussong, 2014; Uljarević et al., 2021). It is conceivable that the low level of metacognitive ability in adolescents (dos Santos Kawata et al., 2021) could lead to poor self-esteem as well as inaccurate confidence over one’s actions and decisions (i.e., continued substance use) regardless of previous negative outcomes (Hauser et al., 2017). Furthermore, the friendship network and the quality of relations between peers could increase the risk of SUD through inducing negative affect (i.e., bullying relationships) or encouraging substance use as a norm and value of the group (Shadur & Hussong, 2014). Family relationship variables (e.g., having deviant siblings, parent warmth) are another group of risk factors that potentially affect adolescents’ substance use initiation (Neiderhiser et al., 2013, Slesnick et al., 2002). Based on these findings, we may postulate that social factors in terms of social stress and social learning process, could act differently across individuals due to their differences in brain structures, that make some adolescents more prone to SUDs. Therefore, low levels of self and social awareness could affect the ability to regulate one’s behavior within a social context.

These findings suggest how RDoC domains/constructs could potentially contribute to the emergence of SUD in adolescents, and in turn, may respond to prevention interventions in terms of neural and behavioral alterations. In the following section, we provide a summary of possible approaches that could possibly target these domains/constructs in order to reduce the risk of SUD.

## CLASSIFICATION OF THE PREVENTIVE APPROACHES BASED ON THE RDoC DOMAINS

One of the main motivations for developing the RDoC framework is to provide an opportunity to link an assessment or intervention in one unit (level) of analysis to other units (Dell’Acqua et al., 2023. As an example, you can start with a behavioral assessment or intervention that targets the positive valence domain and connect it to the corresponding cellular and molecular mechanisms or corresponding neural circuits (Upadhyay et al., 2022). This also makes it possible to integrate interventions that target the same domain/sub-domains at different levels with a hope that our mechanistic understanding will provide synergistic effects and better outcomes.

Based on the RDoC framework and quantitative findings from molecular genetic studies, structure and functional brain research, studies investigating the influence of environmental factors (e.g., poverty, loneliness), and studies investigations of psychological processes, the interventions can be divided into at least 4 major categories (Dalgleish et al., 2020; Rezapour et al., 2017). (1)

Medications and other molecular interventions (e.g., gene therapy) which mainly target Genes, Molecules and Cells units of RDoC (Salles et al., 2020; Hui, 2020). (2) Neuromodulation (Brain Stimulation and Neuro and Biofeedback) which targets mainly Circuits and Physiology levels (Cho et al., 2022; Deng et al., 2020). (3) Cognitive and Behavioral Interventions which mainly target Physiology, Behavior and Self Reports units (Alexopoulos & Arean, 2014; Garber & Bradshaw, 2020; McKay & Tolin, 2017; Burkhouse et al., 2023). (4) Environmental Interventions which target the environment in multiple levels of its microsystems (family and school), mesosystems (community and workplace), and macrosystems (society, ethnicity/gender, culture and ecology) (McLaughlin & Gabard-Durnam, 2022). Although most of the currently available preventive interventions target cognitive and behavioral processes and their relevant environmental components, mapping them into the RDoC framework will connect them to other levels of analysis like corresponding neural circuits and chemical messengers (Steffen, 2023) Future trials can plan to use outcome measures in other levels, like EEG or fMRI to assess and predict responders or measure the response not only in the behavioral and self-report levels, but also in the circuit and molecular level. Future trials can also combine interventions on various levels of the same domain/subdomain to synergistically engage the targeted mechanism and improve the outcomes.

The following section provides an overview of the behavioral and cognitive interventions that can be implemented for targeting risk and protective factors identified for substance use disorder within the RDoC framework above.

### Interventions targeting Negative Valence Systems (NVS)

This group of interventions broadly includes a set of educational and practical techniques termed as “*Emotion Regulation (ER)*”, which are applied to manage negative emotions and their expression in the face of emotional situations, specifically when decision making is required (Hadley et al., 2019). ER encompasses a broad range of skills delivered through emotion education (e.g., identifying triggers, recognizing and labeling feelings) and strategy teaching, including distraction, self-expression, physical exercise, and cognitive reappraisal (Houck, Hadley, et al., 2016). It is worth noting that affective valences are critically associated with somatic cues. Most of the developed ER programs affect the ARS and SP domains as well (e.g., mind–body practices), through increasing individuals’ awareness about their interoceptive signals and emotional states in the face of arousal-eliciting situations.

### Interventions targeting Positive Valence Systems (PVS)

Interventions in this group are largely intended to interfere with an individual’s preference toward immediate rewards (e.g., substance use) and enhance the valuation of delayed rewards (e.g., college graduation). Therefore, preventive interventions which target delay discounting and reward sensitivity through expanding adolescents’ temporal window could potentially reduce drinking alcohol or using substances (Dennhardt et al., 2015).

Moreover, some interventions such as “*Behavioral activation*” could be implemented to increase the rewarding properties of substance-free activities and encourage individuals to engage in these activities on a daily basis (Reynolds et al., 2011). During the course of behavioral activation, individuals are asked to identify their life goals/values and track the enjoyable activities they do in line with these goals/values (Reynolds et al., 2011). The PAX Good Behavior Game is a sample approach which have been developed to encourage prosocial behaviors (e.g., reducing drinking alcohol) through creating a shared relational network of prosocial behaviors, assigning positive value to them and reinforcing one’s engagement (Johansson et al., 2020).

Another group of interventions that is likely to adjust the PVS are educational programs developed with the aims of leveraging individuals’ knowledge about substances and providing them with a perspective on the harms and costs of using substances as well as emphasizing the importance of health as a life value to accomplish long-term goals (Debenham et al., 2020, Sussman et al., 2002). Therefore, the gained knowledge may be able to interfere with reward valuation and expectancy regarding substance use. It is noteworthy that the traditional addiction preventive education programs have recently undergone subtle changes in their content and structure. As a result of this transition, a new concept of “Neuroscience-based Psychoeducation” has emerged, which has been used to convey harm-minimization information to adolescents (Debenham et al., 2020; Ekhtiari et al., 2017). The *Illicit Project* and the *Just Say Know* programs are samples of the pioneers in this field developed to improve adolescents’ neuroscience-based substance literacy level (Meredith et al., 2021; Debenham et al., 2020).

### Interventions targeting Cognitive Systems (CS)

This group of interventions includes all those approaches that tend to promote forethoughtful, and goal-oriented behaviors in which a person could mentally reflect on the consequences of their potential choices. This category mainly relies on a set of processes from basic to more complex cognitive functions activated through using cognitive training and knowledge development. Cognitive training is among the most common components of these interventions, traditionally provided in terms of cognitive games. For example, some studies examined the efficacy of such games (including Lumosity, City Builder game, Fling game) targeting executive functions (e.g., working memory, response inhibition) within a training context (Boendermaker et al., 2017, 2018; Mewton et al., 2020).

Interestingly, the CS could also be targeted by multi-component interventions such as Life Skills Training (Griffin et al., 2006a), RealTeen (Schwinn et al., 2016), and Preventure (Conrad, 2016) which enhance personal competence in terms of complex cognitive skills (e.g., goal setting, planning, decision-making, self-monitoring, problem solving) to motivate and facilitate behavioral change and resist against risky situations. It is note-worthy that some techniques could target multiple interacting domains; for example, self-monitoring techniques that could be used to increase both self-awareness (SP) as well as self-management skills (CS).

### Interventions targeting Arousal and Regulatory Systems (ARS)

Interventions in this group mainly include approaches to resolve sleep problems and adjust circadian rhythms. Sleep problems are multi-cause conditions, which tend to benefit from multi-component interventions. Broadly speaking, sleep education (e.g., teaching sleep hygiene), sleep monitoring (e.g., recording sleep diary and identifying sleep problems), cognitive strategies (e.g., changing sleep-disruptive thoughts), stress management and relaxation techniques (e.g., diaphragmatic breathing) are among the most common ingredients of sleep interventions used as substance use prevention efforts for adolescents (Dong et al., 2020; Fucito et al., 2017, 2021; Miller et al., 2020; Werner-Seidler et al., 2019). Examples of such multi-component interventions are mind-body practices (including yoga and meditations) developed and applied for at-risk adolescents (Butzer et al., 2017). Although all these interventions are focused on sleep and circadian rhythms, they may also alleviate negative emotions and improve mood and cognitive control.

### Interventions targeting Social Processes (SP)

Interventions in this group are divided into two categories, including interventions that target self-awareness and those that enhance social processing. The former category includes interventions that improve self-knowledge and self-esteem (e.g., resistance skills) and those that work on internal attention to calm the mind, body, and behavior and self-awareness (e.g., mindfulness programs)(Waedel et al., 2020); and educational programs which provide scientific knowledge about the effects of different substances on the brain. These educational interventions, such as the Just Say Know (Meredith et al., 2021), translate neuroscience into understandable content, explaining how substances may change brain structures and function and lead to risky behaviors and SUDs. These informative programs increase individuals’ self-knowledge and insight and provide scientific evidence for why adolescents are more vulnerable to initiate substance use to reinforce self-agency to regulate their own behaviors.

Another group of interventions targeting self-awareness provides feedback and normative information which indicates deviation of one’s behavior (i.e., amount of drinking and cannabis use) from the peer norms (Geisner et al., 2007; Larimer & Cronce, 2002; Pischke et al., 2021b; Riggs et al., 2018). The second category encompasses interventions that teach social skills (e.g., communication skills, developing healthy relationships (Griffin et al., 2006a). Life Skills Training (Griffin et al., 2006b), Unplugged (Faggiano et al., 2010), the Climate Schools program (Newton et al, 2022; Newton et al; 2020) are some well-known examples of preventive programs developed based on social theoretical models and focus on social competence in adolescents.

Using the aforementioned categories of preventive approaches, in the next section we classified the existing evidence-based school-based addiction prevention programs selected through a systematic review. The rationale behind selecting schools is that they are ideal site to offer preventive interventions, since they have a high access to an engaged group of adolescents from diverse backgrounds, which could reduce the affordability and accessibility barriers.

## A SYSTEMATIC REVIEW OF SCHOOL-BASED ADDICTION PREVENTION PROGRAMS FOR ADOLESCENTS

### Search Methods

The systematic review was conducted using the PRISMA framework (Moher et al., 2015). Search terms were selected based on past reviews. PubMed databases was searched using the following search syntax: (“Adolescents” OR “Adolescence” OR “Teens” OR “Teen” OR “College” OR “School” OR “Youth” OR “Youths” OR “Young” OR “Teenager” OR “Teenagers” OR “School” OR “College” [tiab]) AND (“Substance Related Disorder” OR “Drug Use Disorders” OR “Drug Use” OR “Substance Abuse” OR “Substance Dependence” OR “Substance Addiction” OR “Addiction” OR “Drug Dependence” OR “Substance Use Disorder” OR “Drug Consumption” OR “Alcohol Related Disorders” OR “Alcohol Problem” OR “Alcohol Dependence” OR “Alcohol Addiction” OR “Alcohol Abuse” OR “Alcohol Use Disorder” OR “Risky Drinking” OR “Heavy Drinking” OR “Alcohol Use” [tiab]) AND (“Prevent” OR “Preventive” OR “Prevention” OR “Intervene” OR “Program” OR “Intervention”). The search period covered studies published from 1996 to August 2022.

### Eligibility Criteria

In this review, we selected the studies if they met the following conditions:

- Study design: Randomized control trials, written in English and available in full text were included if they also met the below mentioned inclusion criteria:
- Participants: Preventive intervention studies for non-user adolescent students aged 13–18 years were included. Studies of interventions involving families/teachers of students were excluded because these programs have specific components focusing on parent/teacher’s skills and attitude towards adolescents’ addiction, which could not be embedded within the RDoC framework. Moreover, we excluded those studies with students who were clinically diagnosed with a disorder (e.g., attention deficit hyperactivity disorder, depression) or were considered as current alcohol or substance users, or those with alcohol/substance use disorders. Because interventions for these groups have substantial differences.
- Intervention: The intervention had to be introduced as a school-based drug/alcohol preventive approach delivered via face-to-face training or technology (e.g., app, web). Community-based interventions or those conducted in the clinical context (e.g., emergency department) were excluded.
- Outcomes measures. Studies that assessed or were planned to assess (e.g., protocol studies) variables related to drug and alcohol (e.g., knowledge, attitude, intention) were included. Studies were excluded if they were concerned with other negative behaviors (e.g., violence, gambling).

### Data Collection and Analysis

Two independent reviewers (TR, PR) screened each title and abstract per inclusion/exclusion criteria. The selected interventions were coded and reviewed in full text by both reviewers. Using the aforementioned criteria, a total of 22 [unique] interventions out of 101 eligible prevention trials (Table 1, See Figure 4) were extracted and analyzed in terms of the type of intervention developed or applied (the specific term coined for the interventional program) as well as their content explained by their authors (Table 2). Although our initial search yielded various interventions that were successfully implemented for students, such as the Unplugged (Faggiano et al., 2010), the Illicit Project (Debenham et al., 2020), and the Just Say Know programs Meredith et al., 2021, they couldn’t be included in the review. This exclusion was due to their involvement of parents, recruitment of individuals aged >18, or evaluation of effectiveness in a pilot study rather than a randomized trial. We also reviewed <u>Blueprint</u> for the evidence-based programs which aimed to prevent substance use in adolescents. The protocol of this systematic review was registered on the Open Science Framework (OSF) on October 9, 2022: https://osf.io/z4v5m/.

**Table 1.** Selected substance use disorder preventive interventions (n=22) and the provided details about their components.

| **Name of program** | **Detailed components** |
| --- | --- |
| Ready4life (Haug et al., 2021) | Substance-related knowledge and attitudes, normative expectations, and skills for resisting media and peer influences to use, personal self-management (decision-making and problem-solving ability, skills for identifying, analyzing, interpreting, and resisting media influences, skills for coping with negative emotions (e.g., anxiety, anger, and frustration), basic principles of personal behavior change and self-improvement (e.g., goal-setting, self-monitoring, and self-reinforcement) and social skills (communication, initiating social interactions, conversation, complimenting, skills related to male-female relationships, and verbal and nonverbal assertive skills) |
| eCHECKUP TO GO (Dumas et al., 2020) | Personalized normative feedback on peer drinking, positive alcohol beliefs, and positive alcohol expectancies, as perceptions of peer drinking and cognitions about alcohol, protective behavioral strategies (e.g., social activities instead of partying) |
| ALERT, Alerta Alcohol program (Martinez-Montilla et al., 2020) | Change students' beliefs about drug norms and the social, emotional, and physical consequences of using drugs; help them identify and resist pro-drug pressures from parents, peers, the media, and others; build resistance self-efficacy, the belief that one can successfully resist pro-drug influences |
| Kripalu Yoga in the Schools (KYIS) curriculum (Butzer et al., 2017) | Stress management, emotion regulation, self-appreciation, confidence, and strong peer relationships |
| The GOOD life (Stock et al., 2016; Vallentin-Holbech et al., 2018) | Social Norms Intervention (social norm and attitude) |
| Brief alcohol intervention (Giles et al., 2016; 2019) | Personalized feedback about the individual student's drinking behavior and attitude, behavior change counselling, advice about the health and social consequences of continued risky alcohol consumption |
| Preventure (Barrett et al., 2015; Conrad, 2016) | Psycho-education (personality profile), behavioral and cognitive coping skills (e.g., goal setting, cognitive restructuring), motivational interviewing |
| MobileCoach Alcohol (Haug et al., 2014) | Social norm interventions, outcome expectations, self-efficacy, planning processes |
| Saluda program (Hernández-Serrano et al., 2013) | Information on alcohol and illicit drugs, and their effects; information on causal factors (motivations) for drinking and taking pills among adolescents; advertising analysis; social skills and assertiveness skills; information on healthy leisure offers existing in the city; abilities in problem solving and decision making; public commitment to non-abuse of alcohol and synthetic drugs |
| Motivational interviewing-oriented brief alcohol interventions (Gmel et al., 2012) | Knowledge, attitude, motivation |
| Brief intervention for poly-drug use (Arnaud et al., 2012) | Personalized feedback on their substance consumption patterns including the associated risks (related to health and other consequences) and comparisons to a normative reference group. Interactive MI-based exercises that have been proven effective in prompting readiness to change by encouraging the participant to consider the costs and benefits of their current substance use and actual change. Practical advice concerning alternative behavior in tempting situations, with a focus on peer resistance skills to raise self-efficacy beliefs and implementation intentions. |
| Drug education program (Midford et al., 2012) | Knowledge, change of thinking, drug-free activities, norms, analyzing, media and social pressure, risk reduction, personal confidence (e.g., positive self-talk, refusal skills, peer negotiation), decision making, assertion skills |
| Project Toward No Drug Abuse (Sussman et al., 2011) | Active listening and communication skills, Stereotyping and social norms, chemical dependency and consequences of drug use, coping with stress, self-control, being assertive, positive/negative thought and behavior loop, attitude, decision-making |
| IPSY (Spaeth et al., 2010) | Intrapersonal and interpersonal Life Skills (e.g., self-awareness, coping strategies, assertiveness, or communication skills), instruction on substance-specific skills (e.g., resistance to peers offering substances). Knowledge concerning alcohol and tobacco use (e.g., prevalence rates, short-term effects, advertising strategies) |
| Cognition-Motivation-Emotional Intelligence-Resistance Skills (CMER) (Guo et al., 2010) | Knowledge about psychoactive substances, the adverse consequences of these substances, anti-drug attitudes and social norms. Motivational intervention to change intrinsic motivation for addictive behavior, coping skills to enhance overall personal competence and to decrease vulnerability to social influences of drug use. |
| RealTeen (Schwinn et al., 2010) | Goal setting, decision making, coping (particularly with stress, puberty, and bodily changes), self-esteem, assertion, communication, media influences, peer pressure, and drug facts. |
| Climate Schools (now known as OurFutures) (Vogl et al., 2009; Slade et al., 2021) | Information regarding social, psychological, physical, and legal consequences of drug use, social pressure, media influences, alcohol-free social activities, normative education, resistance-skills training |
| Take Charge of Your Life (TCYL) (Sloboda et al., 2009) | Personal, social, and legal risks and consequences involved in the use of substances including tobacco, alcohol, and illicit drugs and those beliefs that "everybody does it" are not congruent with reported usage data from national studies. Life skills such as communication, decision-making, assertiveness and refusal skills. Students learn to make sense of experiences, ideas and values about the health, social, and legal risks and consequences of substance use by comparing them with their existing understandings and beliefs. |
| keepin' it REAL (Kulis et al., 2007) | Knowledge, anti-drug norms and attitudes and to facilitate the development of students' risk assessment, decision-making, and ability to refuse drugs, manage stress, interact effectively. |
| Reconnecting Youth (Hallfors et al., 2006) | Self-esteem, decision making, personal control, and interpersonal communication |
| Life skills training (LST) (Seal, 2006; Botvin et al., 1990) | Information and skills specifically related to drug and tobacco use, such as the effects of drugs, self-awareness skills, decision-making and problem-solving skills, stress and coping skills, and refusal skills. |
| Alcohol Misuse Prevention Study (AMPS) (Shope et al., 2001) | Awareness of the short-term effects of alcohol, risks of alcohol misuse, situations and social pressures to misuse alcohol. Skills for resisting against social pressures, self-agency, |

**Table 2.** Substance use disorder preventive interventions (n=22) based on the targeted RDoC domain (s)

| Name of program | NVS | PVS |  |  | SP |  |  |  |  |  |  |  | CS |  |  | ARS |
| --- | --- | --- | --- | --- | --- | --- | --- | --- | --- | --- | --- | --- | --- | --- | --- | --- |
|  | Skills for coping with negative emotions | Motivation to change behavior (e.g., self-reinforcement) | Drug-free activities | Substance-related knowledge (e.g., consequences) | Substance-related knowledge (e.g., consequences) | Attitude towards substance use | Social norms (popularity and prevalence) | Social pressure resisting skills (e.g., awareness, refusal skills) | Social skills (e.g., communication skills, assertiveness) | Self-related awareness (e.g., personality) and self-efficacy skills | Cognitive Restructuring (way of thinking) | Self-monitoring | Self-management skills (e.g., decision-making, problem solving, analyzing) | Behavior changes and self-improvement skills (e.g., self-monitoring, goal-setting, implementation) | Self-control skills and protective behaviors | Stress management |
| Ready4life | + | + |  | + | + | + | + | + | + |  |  | + | + | + |  |  |
| eCHECKUP TO GO |  |  |  | + | + | + | + |  |  |  |  |  |  |  | + |  |
| ALERT, Alerta Alcohol program |  |  |  | + | + |  | + | + |  | + |  |  |  |  |  |  |
| Kripalu Yoga in the Schools (KYIS) | + |  |  |  |  |  |  |  | + | + |  |  |  |  |  | + |
| The GOOD life |  |  |  |  |  |  | + |  |  | + |  |  |  | + |  |  |
| Brief alcohol intervention |  | + |  | + | + | + | + |  |  |  |  |  |  |  |  |  |
| Preventure |  | + |  |  |  |  |  |  |  | + | + |  |  | + |  |  |
| MobileCoach Alcohol |  |  |  |  |  | + | + |  |  | + |  |  |  | + |  |  |
| Saluda program |  |  | + | + | + |  |  | + | + |  |  |  | + |  |  |  |
| Motivational interviewing-oriented brief alcohol interventions |  | + |  | + | + | + |  |  |  |  |  |  |  |  |  |  |
| Brief intervention for poly-drug use |  | + |  | + | + |  | + | + |  | + |  |  |  | + |  |  |
| Drug education program |  |  | + | + | + |  | + | + | + | + | + |  | + |  |  |  |
| Project Toward No Drug Abuse (TND) |  | + |  | + | + | + | + |  | + |  | + |  | + |  | + | + |
| IPSY | + |  |  | + | + | + | + | + | + | + |  |  | + |  |  | + |
| Cognition-Motivation-Emotional Intelligence-Resistance Skills (CMER) |  | + |  | + | + | + | + | + |  |  |  |  |  |  |  |  |
| RealTeen |  |  |  | + | + |  |  | + | + | + |  |  | + | + |  | + |
| Climate Schools (now known as OurFutures) |  |  | + | + | + |  | + | + |  |  |  |  |  |  |  |  |
| Take Charge of Your Life (TCYL) |  |  |  | + | + | + | + | + | + | + |  |  | + |  |  | + |
| keepin' it REAL |  |  |  | + | + | + | + | + | + |  |  |  | + |  |  | + |
| Reconnecting Youth |  |  |  |  |  |  |  |  | + | + |  |  | + |  | + |  |
| Life skills training (LST) | + |  |  | + | + | + |  | + | + | + |  |  | + |  |  | + |
| Alcohol Misuse Prevention Study (AMPS) |  |  |  | + | + |  |  | + |  | + |  |  |  |  |  |  |

NVS= Negative Valence Systems, PVS= Positive Valence Systems, SP= Social Processes, CS= Cognitive System, ARS= Arousal and Regulatory Systems

Interestingly, most of preventive interventions in our systematic review are multi-component programs having more than one target for intervention and addressing several risk factors for SUD, thereby targeting more than one RDoC domain. For example, one of the best-established prevention programs is the *PreVenture* Program which selectively targets four personality risk factors for SUDs (Conrod, 2016). The traits comprise hopelessness, anxiety sensitivity, impulsivity, and sensation seeking, which are all embedded in this interventional program. Each of the intervention components in the *Preventure* program links to a distinct RDoC domain and has been shown to be associated with risk for specific substance use behaviors and concurrent mental health concerns (Conrod, 2016; Stewart, et al., 2021). For example, sensation seeking, is closely related to PVS domain of the RDoC, and is targeted using psychoeducation, motivational enhancement therapy, and cognitive behavioral therapy techniques specifically focused on reward sensitivity. The impulsivity component of the intervention is relevant to CS and focuses on building motivation and cognitive behavioral skills to help young people manage an impulsive personality style and has been shown to reduce substance misuse as well as risk for conduct disorder symptoms (O’Leary-Barrett et al., 2013). The hopelessness and anxiety sensitivity components are relevant to the NVS domain of the RDoC (although hopelessness might be etiologically related to low PVS and lack of inhibition on NVS). Experimental designs have shown that cognitive-behavioral strategies that differentially target these risk factors show some specificity in reducing risk for substance misuse and clinically significant levels of anxiety disorders and major depression (O’Leary-Barrett et al., 2013). The other example of a multi-dimensional program is life skill programs (LSPs) which target intra-and interpersonal skills (e.g., communication skills, empathy, assertiveness, problem solving and decision-making skills, coping with emotions and stress), as well as training substance-related skills (e.g., resistance skills), changing attitude, and improving substance-related knowledge (e.g., norms) (Wenzel et al., 2009). Therefore, addiction prevention programs such as *LSPs* and *Preventure* could have an integrated approach that targets multiple domains of RDoC for a potentially broader target of intervention.

Broadly speaking, the interventions selected for this review were developed focusing on probable risk factors that have been well-identified in previous studies (Thatcher & Clark, 2008; Hammond et al., 2014). These interventions are provided for adolescents who are inherently assumed to be vulnerable to addiction, irrespective of the core pathophysiological mechanism that may link to addiction. For example, adolescents who may at-risk condition due to their emotional problems (e.g., hopelessness, anxiety) receive the same training as those who are at-risk due to their executive dysfunction. While as we develop a better mechanistic understanding about addiction and its protective and risk factors in the individual level with introduction of new bio and behavioral markers in clinical psychobiology, prevention scientists are moving towards using these mechanistic markers to differentiate vulnerable individuals and thus providing them with more precision and personalized preventive interventions. The RDoC matrix can facilitate this process and connect the previous efforts to develop addiction prevention interventions to the new mechanistic efforts (Figure 3).

**Figure 3.**
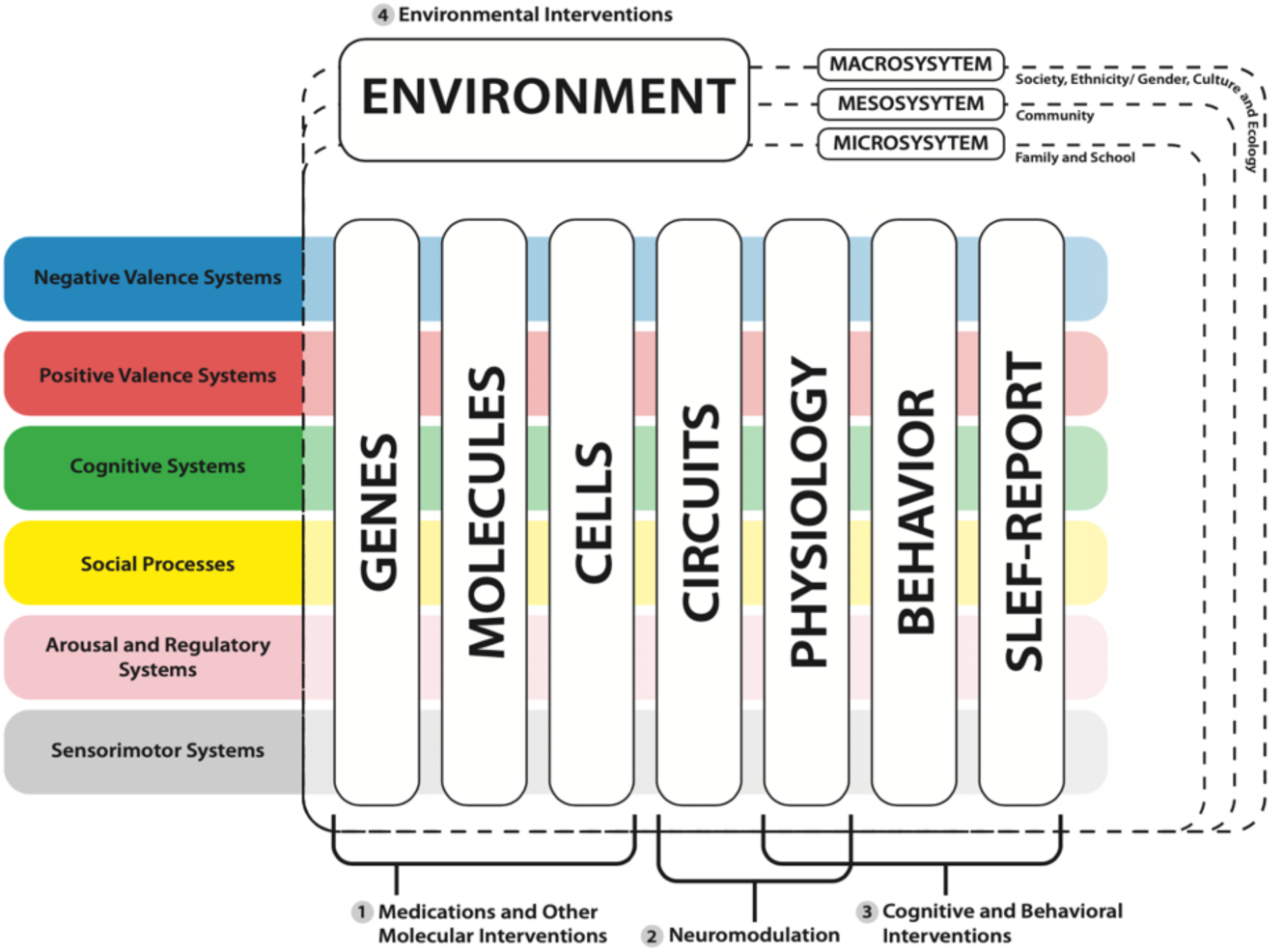
Integration of addiction preventive interventions in multiple domains and levels of analysis in RDoC. While the majority of currently available interventions for addiction prevention are targeting (1) physiology, cognitive and behavioral processes and (2) environment, the RDoC framework provides an opportunity to connect these interventions to other units of analysis like neural circuits or molecular pathways and integrate/combine them with interventions in other levels of analysis. Considering a temporal dimension to the RDoC matrix can provide a developmental perspective to this matrix which is important, especially in preventive interventions for children and adolescents.

**Figure 4:**
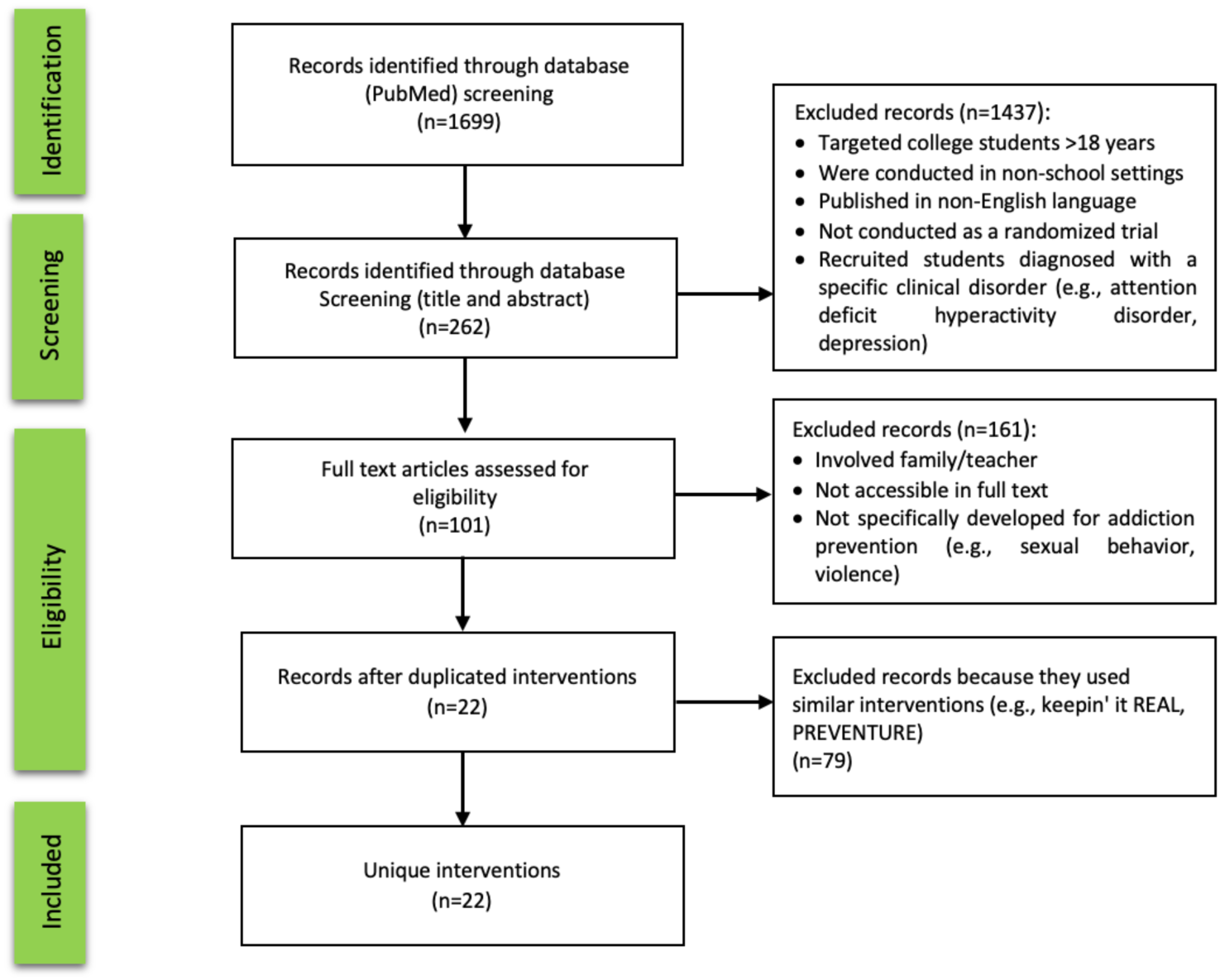
PRISMA summary of identified studies/ interventions included in the review.

## DISCUSSION

In this paper, we described the major RDoC domains involved in SUD and proposed an RDoC-based framework to classify prevention approaches based on their potential functional targets.

Our systematic review yielded 22 unique school-based interventions for addiction prevention that were applied for nonuser adolescents. The results of our subjective classification showed that the Social Processes domain of RDoC was the most frequently targeted construct in the interventions (n=22, 100%), followed by the Positive Valence Systems (n=18, 81.81%), and the Cognitive Systems (n=15, 68.18%). The least frequently targeted constructs were found to be regulatory systems and the negative valence systems (31.81% and 13.63%, respectively). Although this classification shows most of these interventional packages are not only limited to one domain of RDoC, but they cover multiple domains with a hope for a more comprehensive coverage (shotgun approach), the RDoC framework provides an opportunity to tease apart components of the interventional packages and map them into the RDoC domains. Using assessment markers provided within the RDoC matrix in different units of analysis, future interventions may have more focused target (rifle approach) based on the individual needs. Linking the mechanisms to the risk and protective factors of addiction we identified in this paper within the RDoC domains and identifying the specific vulnerability profile at an individual level is an overarching goal of this new line of research that needs to be explored at different levels of analysis defined by the RDoC framework. In the remainder of this section, we argue how using RDoC could bridge the gaps between science and practice to develop more efficient preventive interventions.

Overall, there are several reasons why the classification, development and application of SUD preventive interventions would benefit from the RDoC framework (Insel et al., 2010). First, the RDoC has delineated the major underlying constructs (negative, positive, cognitive, arousal, social) involved in SUD development that could be measured using different levels of analysis, which include molecular, cellular, neural, behavioral, and self-report assessments. At the macro level, researchers within a shared RDoC framework could contribute to increase harmonization and reduce methodological heterogeneity across studies by using a common set of reliable measures, and thus make their results more comparable and compatible with each other. At the micro level, clinical researchers who tend to identify and screen vulnerable individuals, could benefit from these measures to assess the type and the intensity of dysfunction in each domain, and in turn, develop tailored interventions tapping these systems. Referencing this individualized approach to pinpoint the motive(s) for substance use, could result in more phenotypically matched interventions that may increase the likelihood of long-term success.

There are a few pieces of evidence showing great potentials in using specific personal characteristics that moderate SUD vulnerability to predict the responsiveness to the prevention interventions. For example, in a study on a sample of adolescents with and without conduct disorder, the participants with lower neurocognitive skills (i.e., risk taking) achieved less benefits from the component of intervention targeting impulse control, verbal negotiations, problem solving, and cautious decision making (Fishbein et al., 2006). In another study, participants with impulsivity trait responded better to the inhibitory control interventions, while those with sensation seeking trait were more responsive to the interventions that target positive valence system (Conrod, 2016). These moderating effects reminisce of the compensation and magnification hypotheses that account for degree of benefit that people may gain from cognitive stimulation therapy depending on their baseline characteristics (i.e., pre-training level of cognitive alteration) (Carbone et al., 2022). There is still no published study which has applied the RDoC framework to identify high-risk adolescents and examine their responsiveness to an addiction prevention intervention grounded in RDoC-framework.

Second, the RDoC framework provides a set of standardized paradigms which could be efficiently applied for intervention development. The RDoC framework aims to translate the neuroscience-based findings (i.e., precise developmental trajectory) from big datasets such as Adolescent Brain Cognitive Development (ABCD) and HEALthy Brain and Child Development (HBCD) projects to develop preventive interventions and measure their efficacy with proxy neural outcomes (Casey et al., 2018; Jordan et al., 2020). However, cohort studies must begin to incorporate newer designs (e.g., embedded randomized trials, O’Leary-Barrett et al., 2017; Bourque et al., 2016) in order to increase the pace of discovery around promising intervention strategies (Conrod, 2022).

Third, a number of the interventions included in this overview (and possible future interventions) have an impact on a broader spectrum of outcome variables (e.g., suicidal ideation, depression, externalizing symptoms) and can be considered as transdiagnostic interventions (Lynch et al., 2021). The RDoC framework which abandons the traditional diagnostic approach allows for a more systematic exploration of the interplay between these outcome domains and their specific impact on the biopsychosocial processes leading to substance use (Dalgleish et al., 2020). For example, risk factors that increase the likelihood of developing SUDs are primarily studied within individuals who meet diagnostic criteria for SUDs. However, according to the transdiagnostic approach, there is evidence that risk factors for ADHD may also confer the risk for SUDs, as both are associated with hyperactivity-impulsivity symptoms and emotional dysregulation (Anker et al., 2020). In this respect, integration of the RDoC with other empirical models such as the Hierarchical Taxonomy of Psychopathology (HiTOP) that study psychopathological conditions by their signs, symptoms, maladaptive behaviors and traits, may be more efficient for targeting common mechanisms across varied conditions (Michelini et al., 2021).

Additionally, the RDoC framework offers an opportunity to provide drug-related education and training from the lens of neuroscience that is more engaging, non-judgmental, and favorable for the potential end users who would be preventologists, adolescents and their parents.

By using the standardized guidelines derived from such robust findings, modular preventive interventions could be developed using a holistic approach that could be customized to meet the specific needs of individuals, in line with the precision medicine approach (Collins et al, 2007). For example, for adolescents who have experienced various types of childhood trauma (e.g., loss of loved ones, sexual abuse), interventions which emphasize the negative valence (e.g., emotion regulation), regulatory systems (e.g., relaxation) and social processes (e.g., communicating with supportive therapists through conjoint sessions) could be more effective. Finally, by using an RDoC framework, researchers could measure the efficacy of their interventions by using measures which correspond to specific intervention components.

## LIMITATIONS

There are several limitations to the systematic review. Firstly, this review exclusively focused on school-based preventive interventions implemented on non-user students aged between 13-18 years in a randomized trial. Several well-known interventions, such as Unplugged (Faggiano et al., 2010), Fresh Start (Onrust et al., 2018), All Stars (Giles et al., 2010), and Illicit Project (Debenham et al., 2020), were excluded. This exclusion was because they involved parents, included an early adult population (>18 years), were delivered in pilot or feasibility studies, assessed measures other than alcohol or drug-related variables (e.g., sexual behaviors or violence), or were conducted in settings other than schools (e.g., community-based or after-school programs). Secondly, in this review, we attempted to dissect existing preventive interventions based on RDoC domains and constructs. However, we couldn’t differentiate these programs in terms of the strength of the link between an intervention and the corresponding expected domain/construct. For instance, according to Table 2, Ready4life, Kripalu Yoga in the Schools (KYIS), IPSY, and Life Skills Training (LST) were all classified to target the NVS. Still, there is no differentiation between their intensities. The next step would be to provide a measurable scale for evaluating the content of these interventions, enabling a better goodness-of-fit between the interventions and individuals with specific biotypes.

## CONCLUSION

Overall, we suggest that extension in our mechanistic understanding about drug addiction and its prevention and the available framework provided by the neuroscience-informed RDoC matrix, can provide an opportunity to map the previous efforts in developing preventive interventions into a mechanistic map/matrix. We identified protective and risk factors and their relevant interventions for addiction prevention and systematically mapped 22 available preventive interventions into the RDoC domain to show the feasibility. The RDoC framework has a vast potential for informing SUD prevention, particularly in terms of developing mechanism informed preventive interventions and measuring their mechanistic target engagement and efficacy. Although discussing the effectiveness of these interventions is not within the scope of this paper, the proposed conceptual framework provides an insight into how we can develop holistic prevention programs for adolescents by integrating multiple evidence-based paradigms aimed at multiple mechanistic targets. There are several steps ahead for reaching this overarching aim. First, the proposed RDoC domains should receive approval from the global community of addiction prevention experts and achieve their consensus in a Delphi study. This survey can also assess the agreement on the importance of the proposed domains and sub domains to be included in the preventive interventions or if some domains/sub-domains should be prioritized based on multiple factors such as developmental milestones and vulnerabilities. Secondly, the established model should be mapped onto the existing well-established interventions as well as implemented into universal and selective prevention programs to be applied and examined in terms of feasibility and acceptability among target populations. For the final step, randomized clinical and mechanistic studies should be designed to explore the efficacy of the intervention in target engagement and its long-term effects in terms of engaging different units of analysis in the RDoC domains including and most importantly delaying the onset of substance use and reducing the harms (behavioral gold standard outcome). Surely, the active collaboration of the National Institute on Alcohol Abuse and Alcoholism (NIAAA) and the National Institute on Drug Abuse (NIDA) in the US and their counterparts in other countries through allocating funded grants within this framework would be highly effective in taking steps forward and reaching these overarching aims.

## Data Availability

The uploaded manuscript doesn't include any data.

